# Diagnostic Accuracy of GFAP and UCH-L1 for Acute Intracranial Trauma in a Predominantly Blast-Exposed Cohort of Combat Casualties

**DOI:** 10.64898/2026.10.06.26364753

**Authors:** Alisa Sheldon, Yael Rosen Lang, Avital Perry, Anton Peled, Rotem Hershkovitz, Guy Rosenthal, Keren Asraf, Cecilia Moskovitz, Ram Dolman, Dan Dominissini, Yoram Klein, Avinoah Ironi, Avi Epstein, Geoffrey T. Manley, Shachar Shapira, Raquel C. Gardner

**Affiliations:** Joseph Sagol Neuroscience Center, Sheba Medical Center, Ramat Gan, Israel; Department of Neurosurgery, Sheba Medical Center, Ramat Gan, Israel; The Dworman Automated-Mega Laboratory, Sheba Medical Center, Ramat Gan, Israel; Tel Aviv University, Gray Faculty of Medical and Health Sciences, Tel Aviv, Israel; Critical Care Surgery and Trauma Unit, Sheba Medical Center, Ramat Gan, Israel; Emergency Department, Sheba Medical Center, Ramat Gan, Israel; Department of Neurological Surgery, University of California San Francisco, San Francisco, California, USA; Management Wing, Sheba Medical Center, Ramat Gan, Israel; Department of Neurosurgery, Hadassah-Hebrew University Medical Center, Jerusalem, Israel; Hematology Laboratory Unit, Sheba Medical Center, Ramat Gan, Israel

## Abstract

**Importance:** There is an urgent need for objective blood tests to guide triage of suspected traumatic brain injury (TBI) in far-forward combat environments, where blast exposure is common and neuroimaging is frequently unavailable. Blood-based glial fibrillary acidic protein (GFAP) and ubiquitin carboxyl-terminal hydrolase L1 (UCH-L1) are transforming triage of suspected TBI due to their 99% negative predictive value for ruling out acute intracranial trauma in civilian impact TBI. However, accuracy in military blast-dominant TBI is unclear.

**Objective:** To determine accuracy of day-of-injury serum or plasma GFAP and UCH-L1 levels for ruling-out acute intracranial trauma in a sequential real-world cohort of combat casualties.

**Design:** Retrospective cohort study leveraging existing data from a clinical laboratory validation protocol that processed and stored leftover day-of-injury clinical samples from evacuated combat casualties between January 1, 2024, and February 6, 2025 for subsequent analysis for GFAP and UCH-L1 using FDA-cleared assay thresholds.

**Setting:** Single-center, tertiary referral trauma center.

**Participants:** Day-of-injury plasma or serum GFAP and UCH-L1 levels were available for 137 combat casualties. The cohort was 99.3% male with a median age of 21.8 years; 75.2% sustained blast-related injuries. Most (92.7%) had blood collected within 6 hours of injury.

**Exposure:** Serum or plasma GFAP and UCH-L1 measured with Abbott Alinity-i or i-STAT TBI assays. Elevation was defined as GFAP ≥35 pg/mL or UCH-L1 ≥400 pg/mL on Alinity-i, and GFAP ≥30 pg/mL or UCH-L1 ≥360 pg/mL on i-STAT.

**Main Outcomes and Measures:** Accuracy of biomarker elevation for ruling-out intracranial trauma on head CT, including sensitivity, specificity, and negative predictive value (NPV).

**Results:** Among 137 casualties, 83 (60.6%) were biomarker-elevated. Sixteen participants (11.7%) had CT-confirmed intracranial trauma, of whom 15 were biomarker-elevated, yielding 93.8% sensitivity and 98.2% NPV, and 43.8% specificity. Twenty-three participants (16.8%) had intracranial trauma and/or head/face/c-spine fracture; 21 were biomarker-elevated, corresponding to 91.3% sensitivity, 96.3% NPV, and 45.6% specificity.

**Conclusions and Relevance:** In this real-world cohort of blast-dominant combat casualties, day-of-injury GFAP and UCH-L1 demonstrated high NPV and sensitivity for intracranial trauma, with modest specificity. These findings support the utility of GFAP and UCH-L1 to aid early triage and decision-making in blast-dominant military TBI.

## Introduction

Traumatic brain injury (TBI) is a major global health burden and leading cause of morbidity and disability.^1^ TBI is also one of the most common and operationally significant injuries in modern military conflict, where widespread use of explosive weapons results in frequent blast exposure and early diagnosis may be limited by restricted access to neuroimaging.^2–8^

Blast-related injuries involve mechanisms distinct from civilian blunt trauma and frequently co-occur with extracranial injuries, resulting in complex polytrauma that may alter biomarker kinetics and contribute to diagnostic uncertainty in acute TBI evaluation.^2,6,8^ Military personnel often also experience repeated low-level blast during breaching, artillery, and heavy-weapons training, which has been associated with neurocognitive changes, vestibular disturbances, white-matter abnormalities, and altered circulating biomarkers.^9,10^ Among blast-exposed service members, clinical symptoms and signs are often complicated by co-occurring injuries, variable recall of events, and logistical constraints on early assessment.^11–14^

While acute management of TBI has advanced, accurately diagnosing TBI in the hyper-acute period remains challenging, particularly in combat settings.^7,15^ Although intracranial findings on CT^16,17^ and MRI^18^ serve as established diagnostic markers, these tools are unavailable in far-forward environments. This highlights the need for biologically specific tools to improve diagnostic accuracy in the acute evaluation of combat casualties.

Co-occurring psychological trauma, including acute stress reaction and subsequent posttraumatic stress disorder (PTSD), may further complicate symptom-based diagnosis.^8,11,19^

Blood-based biomarkers, Glial Fibrillary Acidic Protein (GFAP) and Ubiquitin Carboxyl-terminal Hydrolase L1 (UCH-L1) are well-validated markers of astroglial and neuronal injury, respectively, and have been shown to aid in evaluation of acute intracranial trauma in civilian emergency department populations.^20–24^ Blood-based GFAP level is associated with TBI severity and long-term functional outcomes.^25^ The FDA cleared the Abbott Alinity-i and i-STAT-Alinity TBI tests based on large multicenter trials demonstrating high sensitivity and negative predictive value (NPV) for ruling out intracranial trauma on CT in civilian adults presenting to the emergency department with GCS 13-15 within 12 hours of head injury.^21,26^ However, their performance has not been evaluated in combat casualties, where blast exposure and polytrauma may affect biomarker concentrations, and is essential for determining their diagnostic utility in operational military settings.

The Israeli operational setting—characterized by small geographic size, proximity of combat zones to advanced trauma centers, and a centralized military evacuation system in which casualties are frequently transported to Level I trauma centers even after relatively minor injuries—provides a unique opportunity to evaluate diagnostic tools for combat-related TBI. Evacuated casualties therefore span a broad spectrum of injury severity; in the present cohort, 21.2% were discharged directly from the ED. This setting enables hospital-based evaluation of biomarker performance while retaining relevance to far-forward triage decisions in other military systems.

## Objectives

This study aims to determine the accuracy of Abbott Alinity-i or Abbott i-STAT Plasma TBI tests of day-of-injury serum or plasma GFAP and UCH-L1 levels for ruling out intracranial trauma in combat casualties presenting within 24 hours of injury. We hypothesized that the assay would demonstrate a 99% NPV for intracranial trauma using established thresholds.

## Methods

### Study Design and Ethics

This is a retrospective single-center cohort study, approved by the Sheba’s Institutional Review Board. The need for informed consent was waived due to the use of existing data generated during routine clinical care and clinical laboratory activities.

### Sources of Data and Inclusion Criteria

Clinical data were obtained from the electronic medical record (EMR). TBI symptom screening was conducted by the Sheba War-Related TBI Task Force when feasible.

Biomarker measurements were performed by the clinical laboratory during validation of the Abbott Alinity-i TBI test from January 2024 through February 2025 using leftover clinical samples, as part of their clinical launch preparation of the FDA-approved test in the Sheba Emergency Department (ED). Inclusion criteria were: (1) emergently evacuated combat casualty arriving to Sheba ED, (2) the clinical laboratory successfully saved leftover serum or plasma from a routine clinical blood collection obtained within 24 hours of injury and subsequently analyzed for GFAP and UCH-L1 levels using the Abbott TBI blood test. All combat casualties arriving at the Sheba ED during the study period had leftover clinical samples saved for analysis regardless of TBI suspicion; exclusions occurred only when samples could not be saved for logistical reasons, including insufficient volume or unavailable staff for timely processing.

### Baseline Characteristics

Blast-related injury was defined as injury associated with an explosive event, without attribution to a specific injury mechanism; such events may involve primary blast overpressure, secondary fragmentation, tertiary bodily displacement or impact, and/or quaternary effects such as burns or inhalational injury.^8^

Baseline characteristics extracted from the medical record included age at injury, gender, Glasgow Coma Scale (GCS) on arrival to the ED, mode of transport to the ED, head CT findings, and hours from injury to blood collection. TBI-related symptoms, including loss of consciousness (LOC), alteration of consciousness (AOC), and post-traumatic amnesia (PTA), were inconsistently documented in the EMR; LOC was therefore coded as present, absent, or not documented.

### Head CT Classification

Participants were categorized as “CT+” or “CT−” based on acute intracranial trauma on the first day-of-injury head CT, as determined by the final clinical radiology report. Those without day-of-injury head CT were coded as “CT not indicated” when the ED trauma team determined that imaging was not clinically indicated. CT-participants were additionally classified for isolated face, skull or cervical spine (c-spine) fracture, determined by the final clinical radiology report, as an additional radiological indicator of high-energy mechanism.

### Blood Biomarker Assays

Blood samples were collected by ED clinical staff via phlebotomy into either yellow-top gel activator tubes (serum) or purple-top (plasma) for clinical analysis. After clinically indicated laboratory testing, serum samples were centrifuged at 4000 rpm for 4 minutes and plasma samples at 2100 rpm for 10 minutes. Specimens analyzed immediately or aliquoted into 300 or 500µL and frozen at −80C for later batch analysis on the Abbott Alinity-I TBI platform.^26^ Two service members had insufficient saved sample for Alinity-i analysis; their plasma was analyzed for GFAP and UCH-L1 using the Abbott i-STAT, a cartridge-based immunoassay requiring 20µL and demonstrating nearly identical accuracy for discriminating acute intracranial trauma on head CT in civilian impact TBI.^27,28^

Alinity-i TBI Test: GFAP and UCH-L1 elevation were defined as concentrations ≥35 pg/mL and ≥400 pg/mL, respectively; participants were classified as blood-elevated if either exceeded threshold and blood-not-elevated if both were below threshold. Both assays used 6-point calibration. GFAP calibration points were 0, 50, 400, 2,000, 20,000, and 50,000 pg/mL; the limit of detection (LoD) and limit of quantitation (LoQ) were 3.2 and 6.1 pg/mL, respectively, with a reportable range of 3.2–42,000 pg/mL. UCH-L1 calibration points were 0, 200, 500, 1,000, 5,000, and 25,000 pg/mL; the LoD and LoQ were 18.3 and 26.3 pg/mL, respectively, with a reportable range of 18.3–25,000 pg/mL. Plasma or serum was used according to available volume after laboratory validation demonstrated near-perfect correlation of GFAP and UCH-L1 concentrations in paired specimens. i-STAT TBI Plasma Test: GFAP and UCH-L1 elevation were defined as concentrations ≥30 pg/mL and ≥360 pg/mL, respectively; participants were classified as blood-elevated if either exceeded threshold and blood-not-elevated if both were below threshold. The LoQ was 23 pg/mL for GFAP and 70 pg/mL for UCH-L1, with reportable ranges of 30–10,000 pg/mL and 200–3,200 pg/mL, respectively. Values outside the reportable range were imputed to the respective lower or upper limits for statistical analyses.

In a paired subset of 42 plasma specimens, GFAP and UCH-L1 concentrations were measured on both platforms to evaluate cross-platform classification agreement using the platform-specific thresholds described above.

### Statistical Analysis

Continuous variables were summarized using medians with interquartile ranges (IQRs), and categorical variables as frequencies and percentages.

Time from injury to blood collection was examined descriptively and displayed in scatterplots but was not used as an adjustment variable. Biomarker concentrations were non-normally distributed and were log-transformed for graphical visualization; nonparametric tests were used for all group comparisons. Diagnostic accuracy metrics—including sensitivity, specificity, NPV, and positive predictive value (PPV)—were calculated using two-by-two contingency tables, treating CT-confirmed intracranial trauma as the reference standard. Agreement in biomarker classification between the Alinity-i and i-STAT platforms was assessed using percent agreement and Cohen κ. Biomarker elevation was defined using the FDA-cleared thresholds for each platform, described above.

Differences in biomarker concentrations across diagnostic categories were assessed using the Kruskal–Wallis test, with Wilcoxon rank-sum tests for pairwise comparisons.

Receiver operating characteristic (ROC) curves and corresponding areas under the curve (AUCs) with 95% confidence intervals were generated for GFAP, UCH-L1, and the combined assay. AUCs were estimated using nonparametric methods.

All statistical tests were two-sided, and p<0.05 was considered statistically significant. No imputation was performed for missing data. Statistical analyses were performed using R statistical software (R Foundation for Statistical Computing, version 4.5.2 [2025-10-31], Vienna, Austria).

## Results

A total of 137 combat casualties met inclusion criteria. The median age was 21.8 years (range 19–43.4), and 99.3% were male, 92% had GCS ≥13 on arrival to our ED, 75.2% sustained blast-related injuries. Overall, 92.7% of blood samples were collected within 6 hours of injury.

Two participants (1.5%) were analyzed using the Abbott i-STAT TBI Plasma assay due to insufficient residual sample volume for analysis on the Abbott Alinity-i platform. One service member presented with GCS 15 and blood collection 2.0 hours post-injury; biomarker concentrations were GFAP 78 pg/mL and UCH-L1 ≥3,200 pg/mL. The second service member presented with GCS 3T and CT-confirmed intracranial trauma, with blood collected 2.3 hours post-injury; biomarker concentrations were GFAP ≥10,000 pg/mL and UCH-L1 ≥3,200 pg/mL. Both participants were classified as biomarker-elevated using the i-STAT assay thresholds.

CT imaging identified 16 casualties with intracranial trauma (IC group) and 7 casualties with isolated fractures of the skull, facial bones, or c-spine (F group). Among the remaining casualties, 88 had complete clinical data; of these, 24 (27.3%) endorsed traumatically induced LOC (LOC group). See **Table 1** for baseline and injury characteristics.

**Table 1.** Baseline Characteristics.

|  | Overall (N=137) |
| --- | --- |
| **Age** |  |
| Median [Min, Max] | 21.8 [19.0, 43.4] |
| **Gender** |  |
| Male | 136 (99.3%) |
| **Mechanism** |  |
| Blast-related | 103 (75.2%) |
| Non-blast-related | 34 (24.8%) |
| **GCS** |  |
| 15 | 124 (90.5%) |
| 13-14 | 2 (1.5%) |
| 3T-12 | 11 (8.0%) |
| **CT** |  |
| Intracranial Trauma | 16 (11.7%) |
| Isolated Fx Face/Skull/C-spine | 7 (5.1%) |
| CT negative | 69 (50.4%) |
| CT not indicated | 45 (32.8%) |
| **Blood Elevated** |  |
| GFAP $\geq 35$ pg/mL | 28 (20.4%) |
| UCH-L1 $\geq 400$ pg/mL | 78 (56.9%) |
| Either Elevated | 83 (60.6%) |
| **LOC** |  |
| Present | 24 (17.5%) |
| Absent | 64 (46.7%) |
| Not documented | 49 (35.8%) |

\*Data are presented as median [minimum, maximum] for continuous variables and number (percentage) for categorical variables. CT findings are categorized as intracranial trauma, isolated skull/facial/cervical spine fracture without intracranial injury, or CT negative/CT not indicated.
\*\* Abbreviations: CT, computed tomography; GCS, Glasgow Coma Scale; LOC, loss of consciousness; GFAP, glial fibrillary acidic protein; UCH-L1, ubiquitin carboxyl-terminal hydrolase L1; C-spine, cervical spine.

In the paired subset of 42 plasma specimens analyzed on both platforms, GFAP classification agreement between Alinity-i and i-STAT was 97.6% (41/42; κ=0.92), while UCH-L1 classification agreement was 83.3% (35/42; κ=0.67) (supplemental eTable 1 and eFigure 1).

Figure 1 illustrates the log-transformed concentrations of GFAP and UCH-L1 by time from injury and CT status. Most samples (76%) were collected within 3 hours. Among CT-positive casualties, three samples collected within 3 hours were elevated for UCH-L1 but not GFAP, whereas one sample collected more than 18 hours was elevated for GFAP but not UCH-L1, demonstrating complementary detection by the two biomarkers across sampling times.

**Figure 1.**
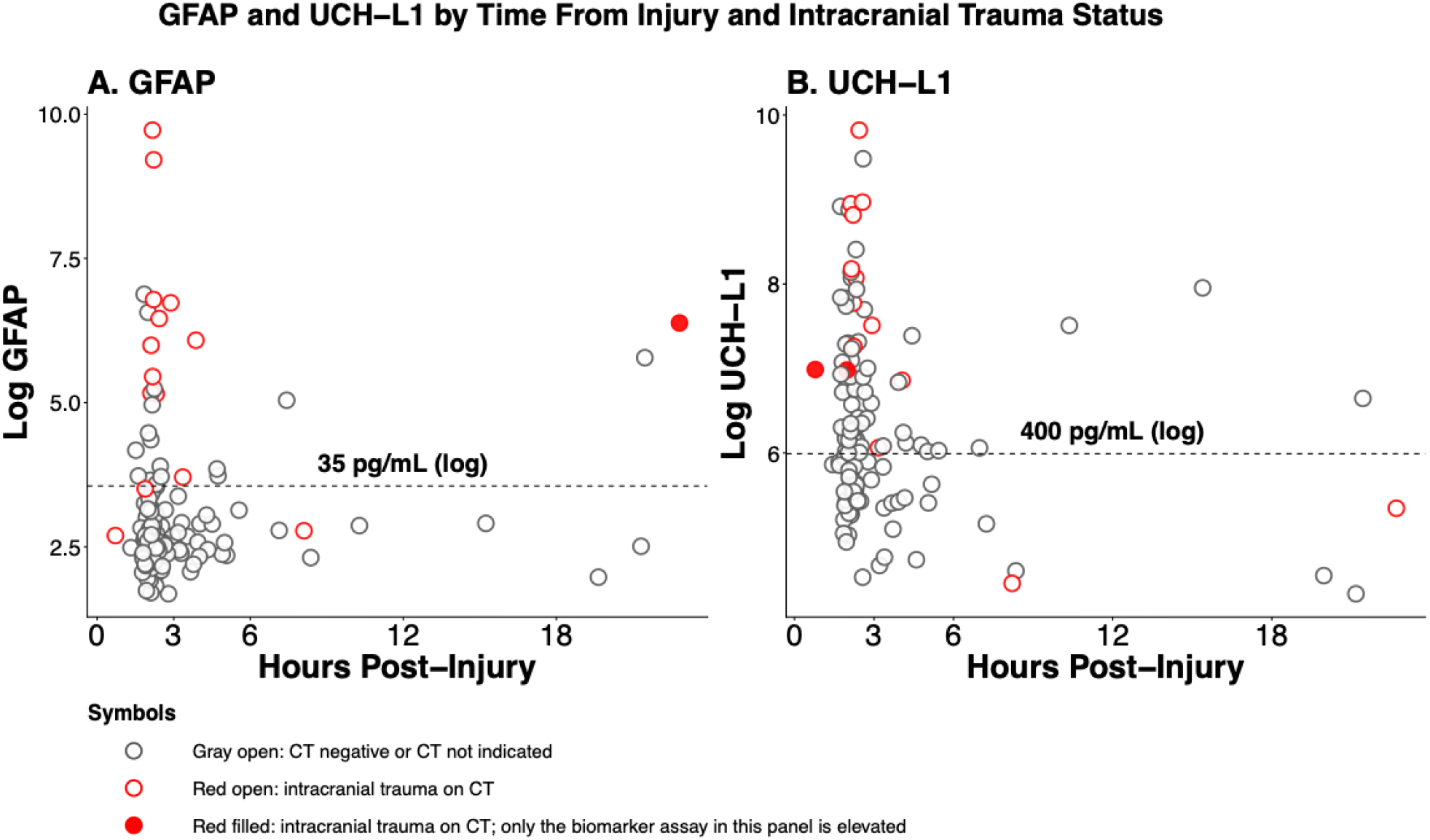
GFAP and UCH-L1 Concentrations by Time From Injury and CT Status. Log-transformed GFAP (A) and UCH-L1 (B) concentrations are plotted against hours post-injury. Gray open circles indicate casualties with a negative CT or for whom CT was not indicated. Red open circles indicate intracranial trauma on CT. Red filled circles indicate intracranial trauma on CT with elevation of only the biomarker shown in that panel. Three samples collected within 3 hours were elevated for UCH-L1 alone; one collected more than 18 hours post-injury was elevated for GFAP alone. Dashed lines mark the biomarker thresholds (GFAP, 35 pg/mL; UCH-L1, 400 pg/mL), shown on the log-transformed axes. Abbreviations: CT, computed tomography; GFAP, glial fibrillary acidic protein; UCH-L1, ubiquitin carboxyl-terminal hydrolase L1.

Using FDA-cleared thresholds (GFAP ≥ 35 pg/mL or UCH-L1 ≥ 400 pg/mL), the combined assay had an NPV of 98%, sensitivity of 94%, and specificity of 44% for CT-positive intracranial trauma; specificity exceeded the 36–37% reported in prior civilian studies.^21^ Among CT-imaged casualties with GCS scores of 13–15, the combined biomarker test had an NPV of 97%, sensitivity of 91%, and specificity of 45%. Among casualties with blast-related injuries within this CT-imaged GCS 13–15 subgroup, NPV was 96%, sensitivity was 90%, and specificity was 45%.

Across the full cohort, 83/137 combat polytraumas (60.6%) were biomarker-elevated. Among these, 15/83 (18.1%) had intracranial trauma identified on head CT. Importantly, 15/16 individuals with CT-confirmed intracranial trauma (93.8%) were biomarker-elevated, demonstrating strong test sensitivity in the subgroup with definitive structural injury. Among biomarker-elevated, CT-negative service members, a substantial proportion endorsed traumatically induced TBI symptoms based on self-report or observer-report, whereas symptom endorsement was markedly lower among biomarker-negative individuals (**Figure 2**).

**Figure 2.**
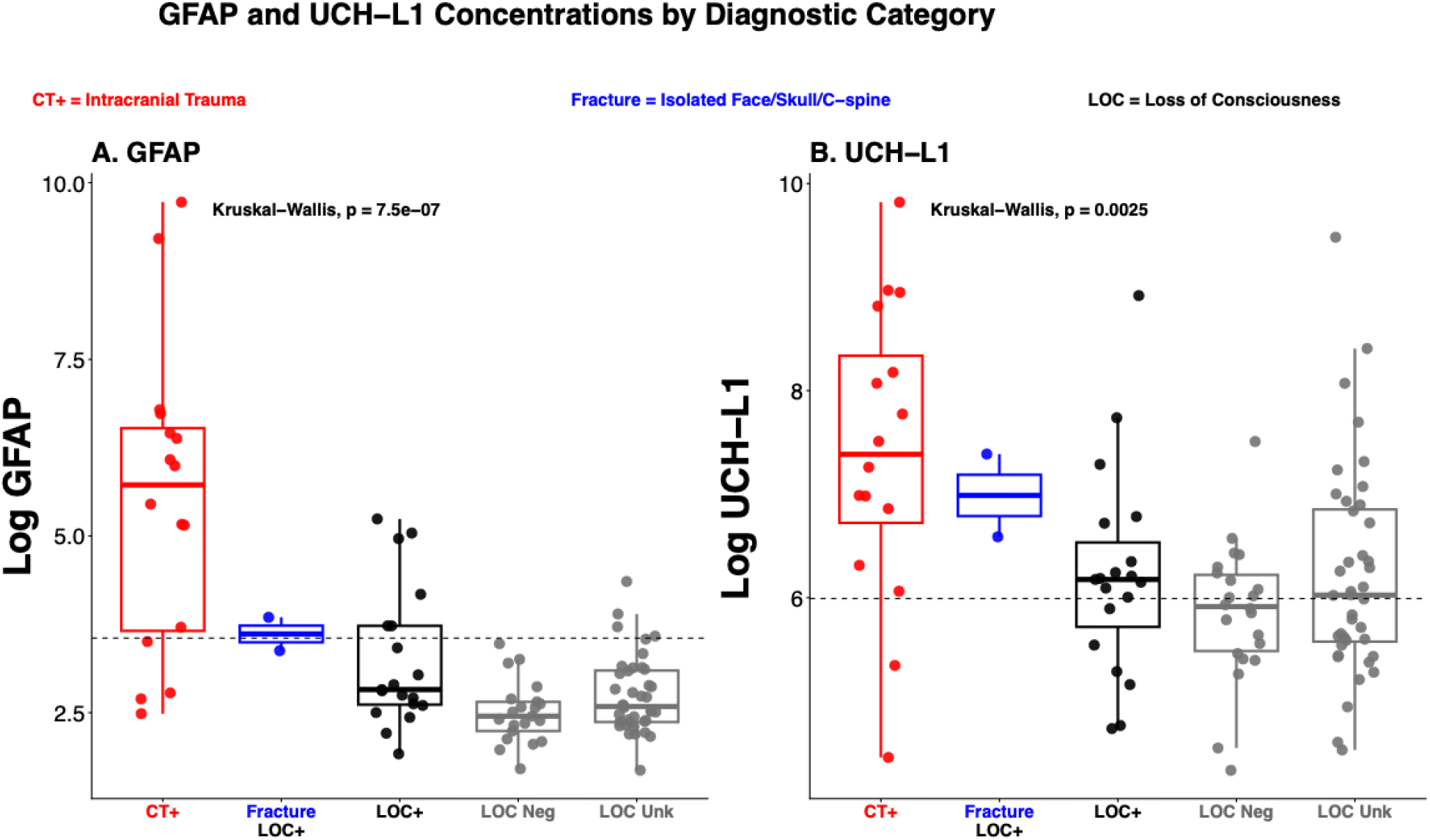
Day-of-injury blood-based GFAP and UCH-L1 levels by CT and clinical features. Boxplots show log-transformed concentrations of glial fibrillary acidic protein (GFAP) (left) and ubiquitin carboxyl-terminal hydrolase L1 (UCH-L1) (right) measured on the day of injury across CT and clinical features: CT+ (intracranial trauma on head CT); Fracture (CT-negative with isolated skull, facial, or cervical spine fracture); LOC+ (CT-negative for intracranial trauma and fracture with reported loss of consciousness without fracture); LOC Neg (CT-negative for intracranial trauma and fracture without reported loss of consciousness); and LOC Unk (CT-negative for intracranial trauma and fracture with undocumented LOC status). Each point represents an individual casualty. Colors denote diagnostic groups (red, CT+; blue, fracture; black/gray, CT-negative groups without fracture stratified by LOC status). Boxes indicate the interquartile range (IQR), with the median shown as the horizontal line within each box; whiskers extend to 1.5× IQR. Horizontal dashed lines indicate assay thresholds (GFAP, 35 pg/mL; UCH-L1, 400 pg/mL). Group differences were assessed using the Kruskal–Wallis test (GFAP p = 7.5×10^−7^; UCH-L1 p = 0.0025).

When stratified by diagnostic category (CT+, F, LOC), GFAP concentrations were highest in the CT-positive intracranial trauma group with minimal overlap with other groups. UCH-L1 concentrations were also elevated in the intracranial trauma group but demonstrated greater overlap with isolated fracture and loss-of-consciousness groups. Both biomarkers were lower in CT-negative casualties (**Figure 2**).

In ROC analyses, the combined assay of GFAP and UCH-L1 demonstrated strong diagnostic performance, with an AUC of 0.87 for intracranial trauma and 0.86 for isolated fractures of the skull, facial bones, or c-spine. When intracranial trauma and fractures were combined as a single diagnostic category (CT+ and/or fracture vs CT−/CT not indicated), the assay yielded an AUC of 0.87 (**Table 2**).

**Table 2.** Diagnostic Performance (AUC) of GFAP, UCH-L1, and the Combined Abbott Alinity-I TBI Blood Test Across Diagnostic Categories.

|  | CT+ (n=16) vs Isolated Fx/CT-/CTNI (n=121)<br>N=137 | Isolated Fx (n=7) vs. CT-/CT<sup>NI</sup> (n=114)<br>N=121 | CT+ and/or Fx (n=23) vs CT-/CT<sup>NI</sup> (N=114)<br>N=137 |
| --- | --- | --- | --- |
| **GFAP** | 0.883 (0.787-0.979) | 0.843 (0.680-1.000) | 0.881 (0.799-0.963) |
| **UCH-L1** | 0.771 (0.619-0.923) | 0.823 (0.674-0.973) | 0.797 (0.680-0.913) |
| **Combined GFAP & UCH-L1** | 0.865 (0.740-0.989) | 0.862 (0.703-1.000) | 0.874 (0.776-0.971) |

\* Values are area under the receiver operating characteristic curve (AUC) with 95% confidence intervals.
\*\* Abbreviations: AUC, area under the curve; CI, confidence interval; CT, computed tomography. CT+: indicates intracranial trauma; CT-: CT Negative; CT<sup>NI</sup>= CT Not Indicated; Fx: isolated skull, facial, or cervical spine fracture.

One false-negative result was observed. This casualty was injured by a single piece of shrapnel from a loitering-munition drone that lodged into his right frontal bone. He presented with GCS 15, and reported not feeling any blast wave, no LOC, PTA, or AOC. Blood was collected 8.25 hours post-injury, with biomarker concentrations below threshold (GFAP 16.1 pg/mL; UCH-L1 86.2 pg/mL). Head CT showed a small focal subarachnoid hemorrhage underlying the frontal bone shrapnel (**Figure 3**). He remained clinically stable and was discharged home without neurosurgical intervention after 7 days of observation.

**Figure 3.**
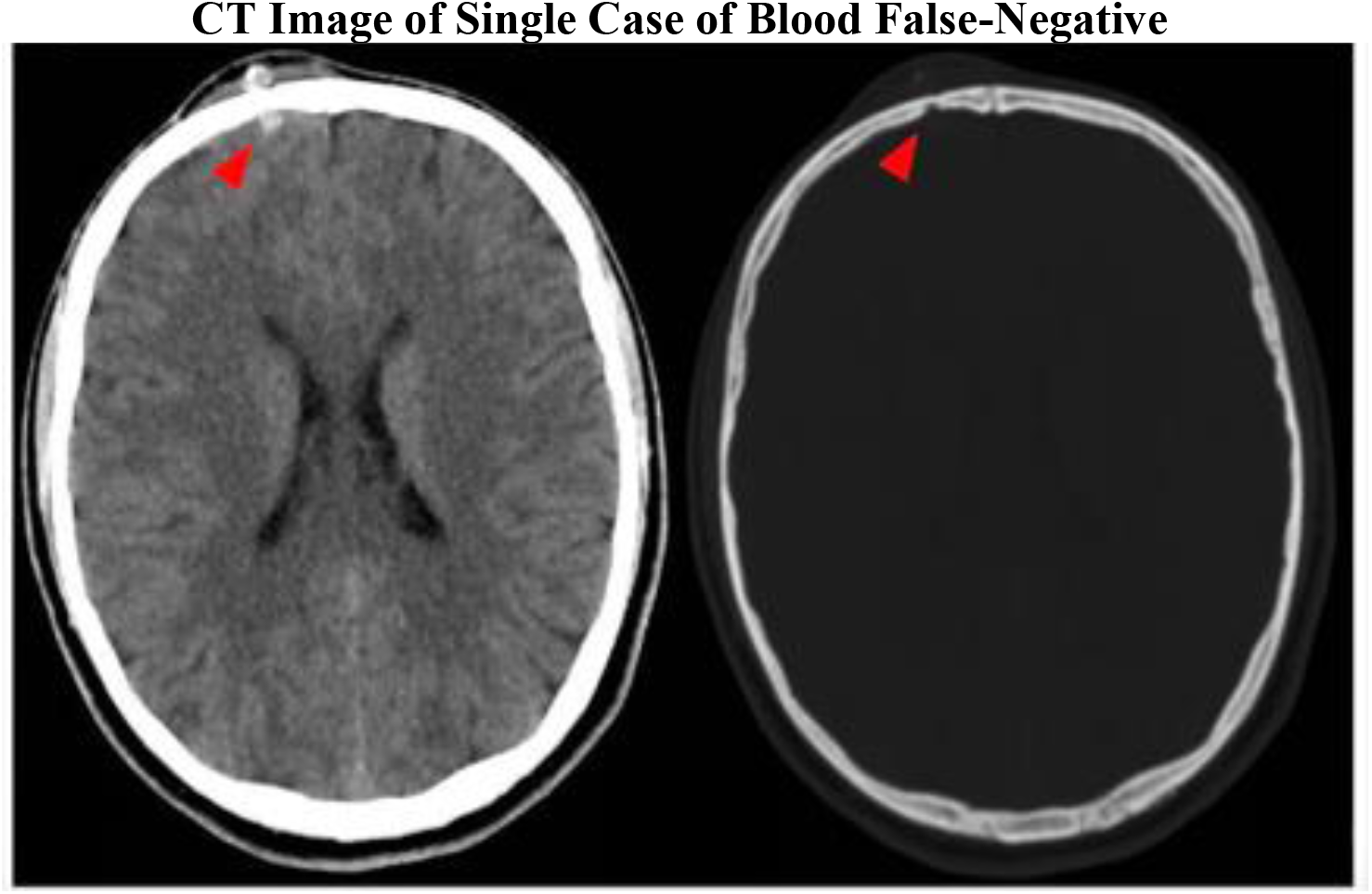
CT Image of Single Case of Intracranial Trauma Without Elevated blood-based GFAP or UCHL1. Representative axial head CT images from the single casualty with CT-confirmed intracranial trauma and GFAP and UCH-L1 concentrations below the Abbott Alinity-i TBI assay thresholds. The brain-window image (left) demonstrates a small focal subarachnoid hemorrhage underlying the right frontal bone injury; the bone-window image (right) demonstrates the corresponding frontal bone shrapnel injury. Red arrowheads indicate the findings.

## Discussion

In this retrospective cohort study of 137 emergently evacuated combat casualties with blood collected within 24 hours of injury, day-of-injury GFAP and UCH-L1 demonstrated high diagnostic accuracy for ruling out CT-positive intracranial trauma, with a NPV of 98% and sensitivity of 94%. Specificity was modest at 44%, though higher than the 36–37% reported in prior civilian cohorts using FDA-cleared thresholds.

### Diagnostic Utility

Our findings align with prior civilian studies evaluating GFAP and UCH-L1 for acute TBI diagnosis. The pivotal ALERT-TBI trial demonstrated a 97.6% sensitivity and a 99.6% NPV for absence of intracranial trauma on CT using these biomarkers, supporting their value as rule-out tools in emergency settings.^21^ Additional analyses from CENTER-TBI and TRACK-TBI similarly showed strong associations between acute GFAP and UCH-L1 levels, CT abnormalities, and injury severity across diverse civilian mechanisms.^20,22,25,29^ Although specificity in our combat cohort (44%) remained modest, it exceeded that reported in earlier civilian studies (36–37%),^21,23^ suggesting robust differentiation of CT-positive and CT-negative outcomes despite heterogeneous battlefield trauma mechanisms.

Interpretation of these results must consider blast exposure and complex polytrauma relative to civilian blunt injuries. Blast-related injury frequently involves extracranial trauma that may influence biomarker levels, yet GFAP and UCH-L1 retained strong discrimination for intracranial trauma despite co-morbid extracranial trauma.^13,14,23,30,31,^ Prior military neuroimaging studies have demonstrated microvascular injury and diffuse axonal disruption in blast-exposed service members despite normal CT findings,^32,33^ underscoring the operational value of objective biomarkers in environments where routine imaging has limited sensitivity for diffuse injury.

In our cohort, 60.6% were biomarker-elevated despite only 11.7% demonstrating CT-positive intracranial trauma, with overlap across isolated fracture, and LOC subgroups. Similar biological injury patterns despite normal CT have been reported in military blast-exposed cohorts using advanced neuroimaging,^32,33^ in repeated low-level blast populations including military and law-enforcement personnel,^9,10^ and in large civilian biomarker validation studies.^21,25^ Repeated low-level blast has been associated with neurocognitive, vestibular, white-matter, and biomarker changes ^9,10^ indicating biological effects without overt TBI. Epidemiological studies have reported that blast-related mTBI in deployed service members is frequently complicated by psychological trauma and heterogeneous injury mechanisms.^7,11,15^ The combination of acute biomechanical brain injury with psychological stress responses contributes to diagnostic uncertainty in CT-negative cases, highlighting the need for biomarkers reflecting underlying neurobiological injury rather than symptom presentation alone.^11,34,35^ Biomarker elevations have also been associated with extracranial trauma, including fractures and spinal injuries,^23,31^ which may partly explain observed overlap between diagnostic subgroups in our cohort. While GFAP demonstrated relatively good separation of CT-positive intracranial trauma from other groups, UCH-L1 showed greater overlap, supporting the use of a combined GFAP and UCH-L1 assay to capture complementary information about acute intracranial trauma.

Strengths of this study include the use of an FDA-cleared biomarker assay in a hyper-acute cohort of emergently evacuated combat casualties, enabling evaluation of GFAP and UCH-L1 performance in a blast-dominant, military environment where such data are limited. Most samples were collected within three hours of injury, providing a rare opportunity to examine early biomarker kinetics under operational conditions. Pre-analytic handling and biomarker analysis methods aligned with expert consensus guidelines.^36^ Clinically indicated head CT enabled classification of intracranial trauma and isolated fractures. Clearly defined diagnostic subgroups enabled comparisons across clinically relevant phenotypes. Collectively, these elements provide a rigorous assessment of biomarker performance in a military population not previously represented in biomarker validation studies.

Limitations include the small sample size, particularly within diagnostic subgroups, and incomplete symptom data for some CT-negative casualties. CT was performed only when clinically indicated, potentially missing intracranial injuries in non-imaged casualties. Biomarker measurements were performed predominantly on Alinity-i, with 2 cases analyzed on i-STAT due to limited residual volume; paired cross-platform testing demonstrated excellent GFAP and substantial UCH-L1 classification agreement. The cohort consisted almost exclusively of male service members, reducing generalizability to women. Finally, while CT provides the operational reference standard, it may not detect diffuse axonal injury or microvascular pathology, frequently observed in blast-related mTBI.^18,32,33^

Future research should include larger, prospectively enrolled military cohorts with standardized symptom assessments and repeated early sampling to refine biomarker kinetics, particularly following blast exposure. Integrating GFAP and UCH-L1 with advanced neuroimaging may establish more sensitive reference standards for CT-negative military TBI and support the development of biomarker-based thresholds specific to CT-negative blast-related TBI, which may differ biologically from CT-negative civilian impact TBI. Further investigation is warranted to determine the prognostic significance of biomarker elevation in isolated skull, facial, or c-spine fractures and CT-negative cases, and to distinguish CT-negative TBI from acute psychological stress responses in combat casualties. Finally, evaluating i-STAT point-of-care use in far-forward settings may inform operational triage and early evacuation decisions.

## Conclusion

In this cohort of emergently evacuated combat casualties, day-of-injury GFAP and UCH-L1 demonstrated strong diagnostic performance for ruling out CT-detectable intracranial trauma, even in a predominantly blast-exposed, polytrauma environment. Early biomarker elevations aligned with clinically relevant injury categories, and the combined assay maintained high sensitivity and NPV despite variable mechanisms of injury. These findings extend prior civilian validation studies by providing the first evidence of biomarker performance in a real-world combat setting and underscore the potential role of GFAP and UCH-L1 as objective, field-relevant tools to support early triage, evacuation prioritization, and clinical decision-making when neuroimaging is limited or unavailable. As military medicine moves toward more rapid, biologically informed approaches to TBI diagnosis, these biomarkers may help bridge critical gaps in care across the continuum from point-of-injury to definitive management.

## Data Availability

The data underlying this study are not publicly available. Requests for access to de-identified data may be directed to the corresponding author and will be considered on a case-by-case basis, subject to approval by the study investigators and institution and applicable ethical restrictions.

## Notes

### Competing Interest Statement

The authors have declared no competing interest.

### Author Declarations

The Institutional Review Board (Helsinki Committee) of Sheba Medical Center, Ramat Gan, Israel, approved this study (protocol No. 9933-22). The committee waived the requirement for informed consent because this retrospective study used existing data generated during routine clinical care and clinical laboratory activities.

