## Supplement for "Diagnostic Accuracy of GFAP and UCH-L1 for Acute Intracranial Trauma in a Predominantly Blast-Exposed Cohort of Combat Casualties"

### Supplement 1. Cross-Platform Comparison of Abbott Alinity-i and i-STAT Plasma TBI Assays

#### eTable 1. Classification Agreement Between Abbott Alinity-i and i-STAT Plasma TBI Assays

| Test | Agreement | Kappa | Sensitivity (Alinity / i-STAT) | Specificity (Alinity / i-STAT) | NPV (Alinity / i-STAT) |
|---|---|---|---|---|---|
| **GFAP** | 41/42 (97.6%) | 0.92 | 80.0% / 80.0% | 89.2% / 91.9% | 97.1% / 97.1% |
| **UCH-L1** | 35/42 (83.3%) | 0.67 | 80.0% / 80.0% | 48.6% / 56.8% | 94.7% / 95.5% |
| **Combined TBI test** | 34/42 (81.0%) | 0.62 | 80.0% / 80.0% | 45.9% / 56.8% | 94.4% / 95.5% |

Agreement was assessed in 42 paired plasma specimens analyzed on both platforms using platform-specific thresholds for biomarker elevation. Abbreviations: GFAP, glial fibrillary acidic protein; UCH-L1, ubiquitin carboxyl-terminal hydrolase L1; κ, Cohen κ.

**eFigure 1. Cross-Platform Comparison of GFAP and UCH-L1 Concentrations Measured Using Abbott Alinity-i and i-STAT Plasma TBI Assays**

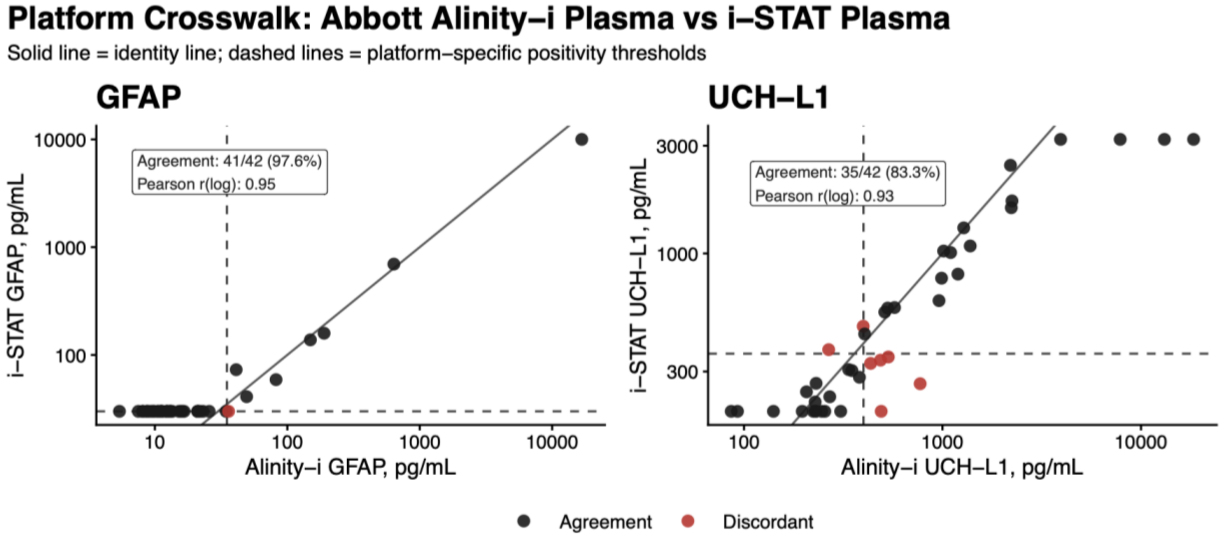


Paired GFAP (A) and UCH-L1 (B) concentrations from 42 plasma specimens analyzed on both platforms. The solid diagonal line indicates perfect agreement between platforms, and dashed lines indicate platform-specific thresholds for biomarker elevation (Alinity-i: GFAP, 35 pg/mL; UCH-L1, 400 pg/mL; i-STAT: GFAP, 30 pg/mL; UCH-L1, 360 pg/mL). Points indicate concordant or discordant classification relative to the platform-specific thresholds. GFAP classification agreement was 97.6% (41/42; κ=0.92), and UCH-L1 classification agreement was 83.3% (35/42; κ=0.67).
